# Nasal microbiota exhibit neither reproducible nor orderly dynamics following rhinoviral infection

**DOI:** 10.1101/2020.04.11.20061911

**Authors:** Sai N. Nimmagadda, Firas S. Midani, Heather Durand, Aspen T. Reese, Caitlin C. Murdoch, Bradley P. Nicholson, Timothy Veldman, Thomas W. Burke, Aimee K. Zaas, Christopher W. Woods, Geoffrey S. Ginsburg, Lawrence A. David

## Abstract

**Background:** How human-associated microbial communities resist and respond to perturbations remains incompletely understood. Viral challenge provides one opportunity to test how human microbiota respond to disturbance.

**Methods:** Using an experimental human rhinovirus infection challenge model, we explored how viral infection may alter microbiota of the upper respiratory tract (URT). Healthy human volunteers were inoculated with HRV serotype 39. Samples were collected by lavage before and after inoculation from healthy (sham inoculated, n=7) and infected (n=15) individuals and subjected to 16S rRNA gene sequencing through amplification of the V4 hypervariable region.

**Results:** No evidence for differences in community alpha-diversity between cohorts was observed. The composition of microbiota of sham-treated and infected subjects did not appear distinguishable and no taxa were significantly associated with infection status. We did not observe support for a correlation between microbial dynamics and counts of specific monocytes. Subject identity was found to be the strongest determinant of community structure in our dataset.

**Conclusions:** Overall, our findings do not suggest a consistent nasopharyngeal microbiota response to rhinovirus challenge. We support the conclusion that this microbial community is individualized. Broadly, our findings contribute to our understanding of how and when immune responses to viruses affect bacterial communities in the URT.

## INTRODUCTION

Longitudinal studies show the composition of human-associated microbial communities (microbiota) can remain steady over long periods of time [1-3]. To understand the forces stabilizing human microbiota, it can be helpful to study processes that disrupt human-associated microbial communities. One such process is pathogen colonization. Pathogen invasion can persistently deplete over half of bacterial taxa in a body site [2]. Concomitant inflammatory responses can also reduce colonization resistance by disrupting indigenous microbial communities [4]. To maintain homeostasis, commensal microbiota may have evolved to resist pathogens by inhibiting the growth or colonization of invading bacteria [5].

Microbial communities residing in the nose and throat are thought to play an important role in pathogen colonization [6]. Acting as the interface between the respiratory system and the environment, the upper respiratory tract (URT) contains a complex network of distinct microbial populations whose distributions are dictated by a number of factors stemming from the environment, the spatial heterogeneity of the URT and host immune function [7, 8]. Less well-understood is how these communities change across infection. Recent work has linked the dynamics of specific microbiota in the URT with susceptibility to infection. For example, an early abundance of *Moraxella and Corynebacterium/Dolosigranulum* in communities was associated with stable dynamics and decreased frequency of reported acute respiratory infections (ARIs) [9, 10]. Shifts towards community dominance by a select few bacteria have been shown to precede the appearance of viral pathogens and symptoms [10]. Moreover, surveys during acute respiratory infections depict a conceivable consistent response to certain viral infections. Studies have found consistent increases or decreases of certain bacterial taxa following influenza disturbance [11-15], and an overrepresentation of *Haemophilus influenzae* has been observed in instances of Respiratory Syncytial virus (RSV) [16-18]. Small amounts of live attenuated influenza vaccine have also repeatedly resulted in increased bacterial diversity up to six weeks post inoculation in the URT [12, 13]. Following these increases, nasal taxonomic diversity decreases over time [18].

Investigating how infections reshape URT microbiota in humans is challenging. Observational studies of human infection often require unique cohorts at high risk of disease [19]. Discovering a predisposing microbiota presents a challenge due to the need for voluntary viral exposure or a long study timeline. Although research to date has already highlighted dynamic, niche-specific communities, it is still in its infancy. No consensus definition of ‘healthy’ or ‘infected’ nasopharyngeal communities in mature adults currently exists in part because many studies have focused on infant or pediatric populations [20].

Here, we address these challenges using a human rhinovirus (HRV) challenge study to examine how viral infection affects the human nasal microbiota. Use of a challenge study design over the course of two weeks enabled us to collect serial microbiota samples of mature hosts before and after infection, as well as control for seasonal effects. Focus on a viral pathogen also let us investigate how host response to infection, and not direct bacterial interactions between a pathogen and commensals, affected human microbiota. Such host immune responses have been associated with URT microbiota shifts including increases in microbial species richness and taxonomic diversity [13, 21]. In the context of viral ARIs, prior work has associated these infections with URT alterations [11-17], and HRV infections specifically have been linked to increases in select genera during infection and a decrease in community diversity [22, 23]. Here, we generate additional observations across 22 participants, to our knowledge forming the largest cohort study of URT microbiota during HRV challenge to date.

## METHODS

### Sample Collection

Healthy volunteers were recruited and challenged with human rhinovirus as previously described [24] under a study protocol approved by the Duke Medicine Institutional Review Board (Durham, NC). Following informed consent and screening, eligible subjects (*n*=30) were enrolled and entered the phase I quarantine facility at Duke’s clinical research unit (DCRU) for 2 days following inoculation with 10^6^ TCID_50_ GMP HRV serotype 39 and subsequent viral challenge [24]. Patients were randomly assigned to challenge and sham groups. Subjects returned for 3 consecutive daily follow up visits through day 5 for daily symptom and sample collections (Figure 1, Table 1). Of the 30 subjects enrolled, 23 completed the study. Of the 23, 7 asymptomatic individuals were inoculated with a sham treatment and 15 exhibited evidence of infection through HRV shedding or seroconversion. One subject (subject 5) was inoculated with rhinovirus but recorded healthy symptoms scores and excluded from analysis for a total of 22 individuals.

**Table 1:** Subject Information.

| Subject | Age, years | Sex | Race | Shedding<sup>B</sup> | Seroconversion<sup>C</sup> | Symptomatic |
| --- | --- | --- | --- | --- | --- | --- |
| 1 | 22 | M | White | - | - | - |
| 2 | 25 | F | White | - | - | - |
| 3 | 27 | F | White | - | Yes | Yes |
| 4 | 35 | M | Black | Yes | - | Yes |
| 5<sup>E</sup> | 24 | M | White | - | - | - |
| 7 | 26 | F | White | Yes | - | Yes |
| 8 | 30 | M | Black | Yes | Yes | Yes |
| 9 | 23 | M | White<sup>A</sup> | Yes | Yes | Yes |
| 10 | 21 | M | White<sup>A</sup> | Yes | - | - |
| 12 | 29 | F | White | Yes | - | Yes |
| 13 | 24 | M | White | - | - | - |
| 14 | 27 | M | Black | Yes | - | Yes |
| 15 | 31 | M | Asian | Yes | - | - |
| 16 | 28 | F | Black | Yes | - | Yes |
| 17 | 24 | M | White | Yes | Yes | Yes |
| 22 | 25 | M | White | Yes | Yes | Yes |
| 23 | 37 | F | American Indian/ Alaskan Native | Yes | Yes | Yes |
| 24 | 29 | M | White<sup>A</sup> | Yes | - | Yes |
| 25 | 21 | M | White | - | - | Yes |
| 27 | 21 | F | White<sup>A</sup> | - | - | - |
| 28 | 40 | M | White | Yes | - | - |
| 29 | 19 | F | Asian | - | - | Yes<sup>D</sup> |
| 30 | 24 | F | White | - | - | Yes<sup>D</sup> |

<sup>A</sup> Hispanic or Latino Specified
<sup>B</sup> Meets criteria for infected/shedding phenotype (measurable viral titer on 2 or more days 24 hours post inoculation)
<sup>C</sup> Convalescent (day 28) Seroconversion
<sup>D</sup> HRV free yet reported symptoms
<sup>E</sup> Excluded from analysis

**Figure 1.**
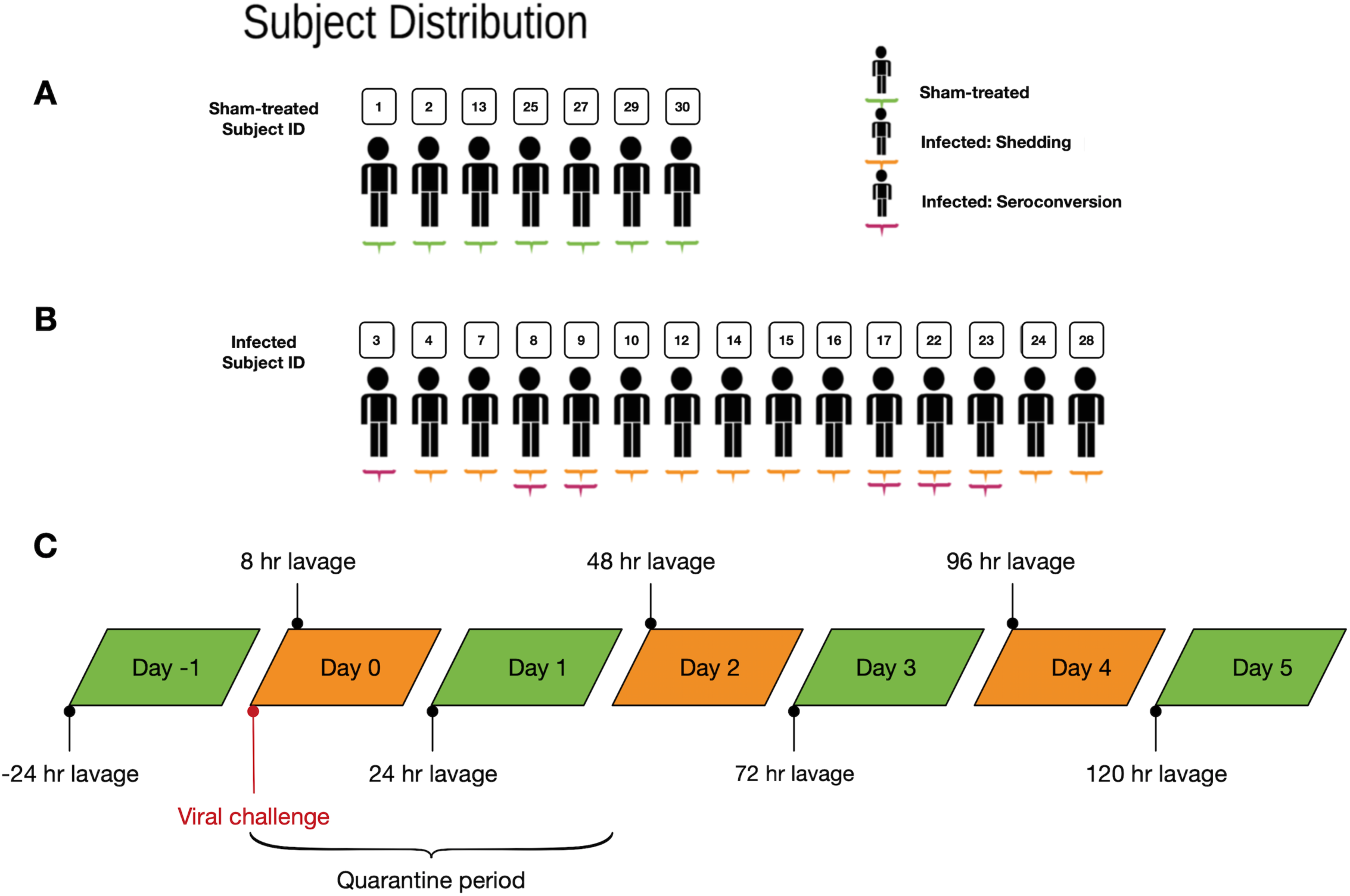
Study design. Subject distribution within the sham-treated (**A**) and infected (**B**) cohorts. Sham-treated subjects are underlined green. Infected subjects exhibiting shedding or seroconversion are underlined in orange and pink respectively. (**C**) Temporal distribution of sampling.

Nasal lavage samples of the URT were collected using 0.9% sterile saline as described [25] at baseline (day-1), post-inoculation (8hr), and daily thereafter (24hr, 48hr, 72hr, 96hr, 120hr). Wash samples were immediately chilled on ice, gently vortexed, aliquoted into cryovials (1mL), frozen and stored at −80°C. These samples were used to determine infection status by quantitative viral cultures. Symptoms were recorded at least twice daily using standardized symptom scoring [26]. We used a modified Jackson Score as published previously [24, 27] to determine those subjects who developed typical cold symptoms of a rhinovirus infection. The highest score per symptom on each day was summed over the course of the challenge (Table S1); the median was used as a cutoff to identify HRV-positive mild and severe response groups.

Prior to inoculation, subjects underwent repeated HRV antibody testing as well as baseline laboratory studies, including complete blood count (CBC) and serum chemistries. Peripheral blood samples from CBC readings were obtained from each subject at predetermined intervals (Table S2).

### DNA Extraction, Amplification, and Amplicon Cleaning

DNA from bacteria was isolated using a MoBio PowerSoil DNA extraction kit. To increase DNA output from nasal lavage fluid (NLF), prior to extraction, we first transferred 800 µl lavage aliquots to a 2 mL tube and centrifuged at 8000 rpm for 20 minutes. DNA extraction then followed the PowerSoil protocol. [28, 29]. The V4 regions of the 16S rRNA gene were amplified from extracted DNA samples utilizing a set of primers with the sequences 5’-CCGGACTACHVGGTWTCTAAT-3’ and 5’-CAAGCAGAAGACGGCATACGAGAT – 3’. All samples were assigned a unique 12 bp barcode and amplified in triplicates along with a negative control. At the end of 35 cycles samples were incubated at 72°C for 10 minutes. PCR products were run on a gel to ensure amplification of the correct size and a lack of contamination. The 16S V4 amplicon was purified and separated using AMPure XP beads (Beckman Coulter Genomics). The purified products (∼33 µl supernatant) were transferred to a fresh 96-well plate and stored at –20°C for later use or quantification.

### Quantification and Sequence Preparation

Initial amplicon quantification was done using the Quant-iT dsDNA High-Sensitivity Assay Kit (Invitrogen). To ensure a sufficient concentration of DNA (20-50 ng/µl) in the sequencing library, amplicons were pooled in ratios ensuring an even representation of DNA from each sample. Pooled products were purified using the MinElute PCR Purification Kit (Qiagen) and gel-purified using the QiaQuick Gel Extraction Kit (Qiagen). Concentration of pooled DNA was verified using the Qubit 2.0 Fluorometer to confirm a minimum volume of 20 µl of pooled sample with a concentration greater than or equal to 20 ng/µl. All pooled samples were sequenced on an Illumina Miseq instrument at the Duke University Sequencing Core using the MiSeq v2 Reagent Kit (Illumina) with paired end, 150 base pair reads. Operational taxonomic units (OTUs) were picked at a 97% sequence similarity threshold using the uclust method referencing the Greengenes dataset (gg_13_8) [30]. OTUs seen in fewer than 5 samples were removed from downstream analysis resulting in 1,982 and 344,982 sequences as the lower and upper bounds respectively (mean 56,685.831, S.D 51,077.356, median 40,948.0).

### Sequence Analysis

Diversity and compositional analysis was undertaken in the QIIME computational environment, versions 1.9.0 and 1.9.1 [31] using phylogenies obtained from the Greenegenes dataset (version 13_8) [30]. Within sample (alpha) diversity of the nasal microbiome was evaluated using counts of observed OTUs, the Shannon diversity index, and the Chao1 index. Analysis of within-population diversity was carried out using a series of scripts in QIIME with a previously published workflow [32]. Between population analysis (beta diversity) was calculated using Bray-Curtis, unweighted Unifrac, and weighted Unifrac distances. PCoA analysis was performed using the principal_coordinates.py and make_2d_plots.py scripts in QIIME.

### Statistical Analysis

To qualitatively and quantitatively compare differences in inter-individual (between subject) and intra-individual (within subject) Unifrac distances, the make_distance_boxplots.py script in QIIME was used. The script performs the students two-sample t-test to identify statistically different distributions; calculation of the non-parametric p-value was done using Monte Carlo permutations. Due to limitations in how implementations of the PERMANOVA calculate effects in a longitudinal study, we split our statistical tests in the following manner. We tested for the effects of Time and an interaction between Time and Infection status using the function call adonis(dm∼Infection*Time + Subject, data=map, strata=Subject permutations=999) where the metadata factor Time signifies time following inoculation (t= −24h, 8h, 24h, 48h, 72h, 96h, 120h). We used the adonis function available in the R (Version 3.2.0) package vegan (version 2.3.0) [33]. To measure the effect of Subject, we reran adonis without Subject as a blocking factor (formula=dm∼Subject, data=map, permutations=999). To test the effect of Infection in light of the nested design of our experiment, we performed a non-parametric ANOVA using the command nested.npmanova(dm∼Infection + Subject, data = map, permutations = 999) available in the R package Biodiversity R (version 2.8-2).

To determine the effect HRV has on community composition in our time series, the Kruskal-Wallis H test was performed at the phylum level (for all phyla) and at the genus level (for common taxa, meaning they were detected in at least 15 samples, resulting in 171 unique genera tested). Relative abundance of individual and clustered taxa (see below) in infected and sham-treated samples was compared at the phyla and genus level using the Mann-Whitney U test with correction for multiple hypothesis testing using the Benjamini-Hochberg method.

Clustering of taxa was performed similarly to a previously published technique [34]. In order to elucidate cluster composition, OTUs were initially grouped at the genus level. 10.7% of taxa were discarded; these taxa were not assigned at the genus level. Rare genera, classified as genera observed in fewer than 5 samples were discarded - rare genera comprised 382 of 647 total observed genera. The remaining 265 genera were included in the analysis and assigned a cluster. The analysis pipeline was as follows: pairwise correlations between genera were first estimated using SparCC [35]. A pairwise dissimilarity matrix was computed as (1-correlation between genera) and was then investigated with the hierarchical clustering toolbox in SciPy version 0.11 [36]. The clustering was carried out using the “linkage” function (method = weighted). Following, taxa were split into clusters utilizing the fcluster function (criterion = distance). Identification of a clustering threshold required a tradeoff between model simplicity and fidelity. Model simplicity entails building clusters with a reasonably interpretable number of genera, whereas fidelity involves capturing the dynamics of more genera. By choosing a clustering threshold of ½ the maximum distance between any 2 genera, we balanced these two ideals. Cluster abundances were reported as the fraction of total 16S rRNA gene reads for a sample.

Counts of leukocytes were correlated with the abundances of six major clusters. To account for potential trends resulting from a comparison of timeseries data, the first difference of each pair was calculated for analysis. Only complete pairs of cluster abundance and monocyte counts were included in the analysis.

## RESULTS

Thirty volunteers were enrolled in the original study [24]. After excluding volunteers with incomplete sampling data and baseline viral contamination, data for twenty-two volunteers was available for sequencing and analysis (Table 1). The average age of participants was 26.73 years. Fifty-nine percent of the subject pool was male and eighteen percent were of Hispanic or Latino origin. Of the twenty-two volunteers, seven were given sham inoculations; these make up the sham-treated subject population (Figure 1). All infected subjects positively reported one or more of the following symptoms common to HRV during the trial: runny nose, stuffy nose, sneezing, cough, malaise, sore throat, headache, shortness of breath, earache (Figure S1). Eight of the fifteen subjects in the infected cohort made up the severe infection group with a sum symptom score greater than 18 over the course of the infected period (Figure S1; Table S3).

A total of 118 samples across 22 subjects were successfully amplified and sequenced, with a representation of all study participants at a minimum of 3 time points (at baseline and at least two other time points). A median of 41,280 reads survived quality filtering and were assigned taxonomy per microbiota sample (min=2,069, max=347,066, median absolute deviation=23,271). Sequences were clustered into 11,462 distinct OTUs (97% identity cutoff). After removing OTUs seen in fewer than 5 samples, 1,784 unique OTUs were left for further analysis. These taxa represented 281 genera from 16 bacterial phyla. A median of 329 unique filtered OTUs were observed in each sample.

We first tested whether species diversity was affected by HRV infection and symptom severity. We computed the alpha-diversity at the OTU level of the nasal microbiota of infected and sham-treated subjects at baseline (Figure S2) and over the HRV exposure period following inoculation (Figure 2). HRV infection was not associated with differences in alpha diversity as measured by the Shannon index, counts of observed OTUs, or Chao1 diversity index (p>0.05, Wilcoxon rank-sum; Figure 2). Within the infected cohort, symptom severity was also not associated with alpha diversity (p>0.05, Wilcoxon rank-sum; Figure S3)

**Figure 2.**
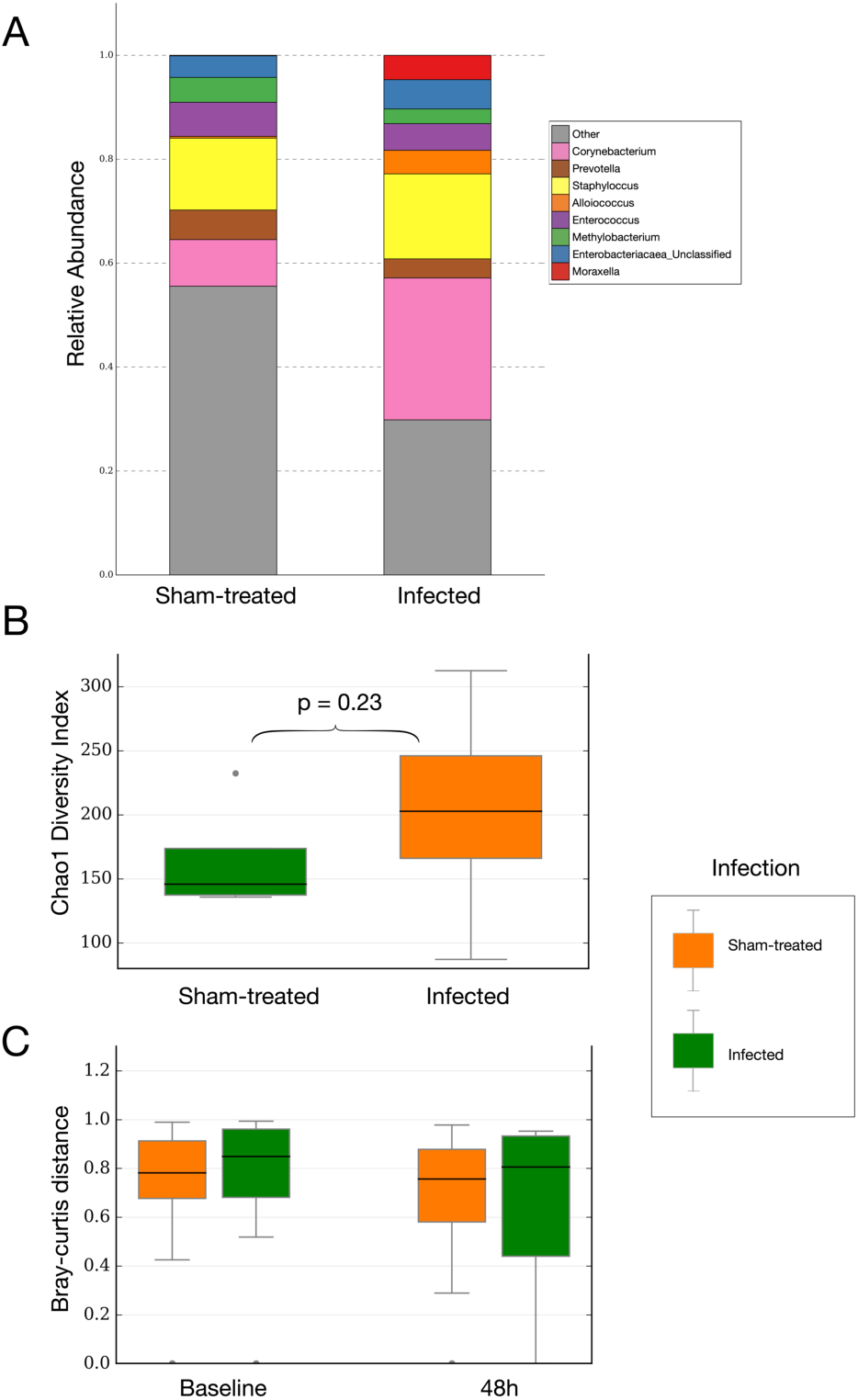
Taxonomic composition of the sham-treated and infected cohorts. (**A**) Stacked bar chart of the median abundance of most abundant genera within the two cohorts during infection at t=48h (p > 0.05; Wilcoxon rank-sum). According to the Greengenes database, operational taxonomic units were considered “unclassified” if the RDP classifier could not assign taxonomy at the family level. Genera comprising less than 1% of the total abundance were combined into the “Other” category. (**B**) Box and whisker plot of Chao1 diversity index. Sham-treated (n=5) versus infected (n=12). No significant differences in alpha diversity between the two cohorts were found (p > 0.05; Wilcoxon rank-sum) (see figure S2 for distributions of baseline alpha diversity and figure S3 for rarefaction curve). **(C)** Box and whisker plot of Bray-Curtis dissimilarity at baseline and 48h.

We next examined whether there were compositional differences associated with HRV infection across the subject pool. We performed principal coordinate analysis (PCoA) of pairwise distances between samples to visualize the similarity of nasal microbial community samples. PCoA plots using on Bray-Curtis, weighted Unifrac, and unweighted Unifrac measures visually clustered by individual identity (Figure S4). By contrast, we did not observe distinctions between samples according to subject health status (Figure 3). Samples also did not appear to be structured temporally among infected patients before, during, and after infection (Figure S4). Similarly, samples did not appear to group by HRV response severity (Figure S5).

**Figure 3.**
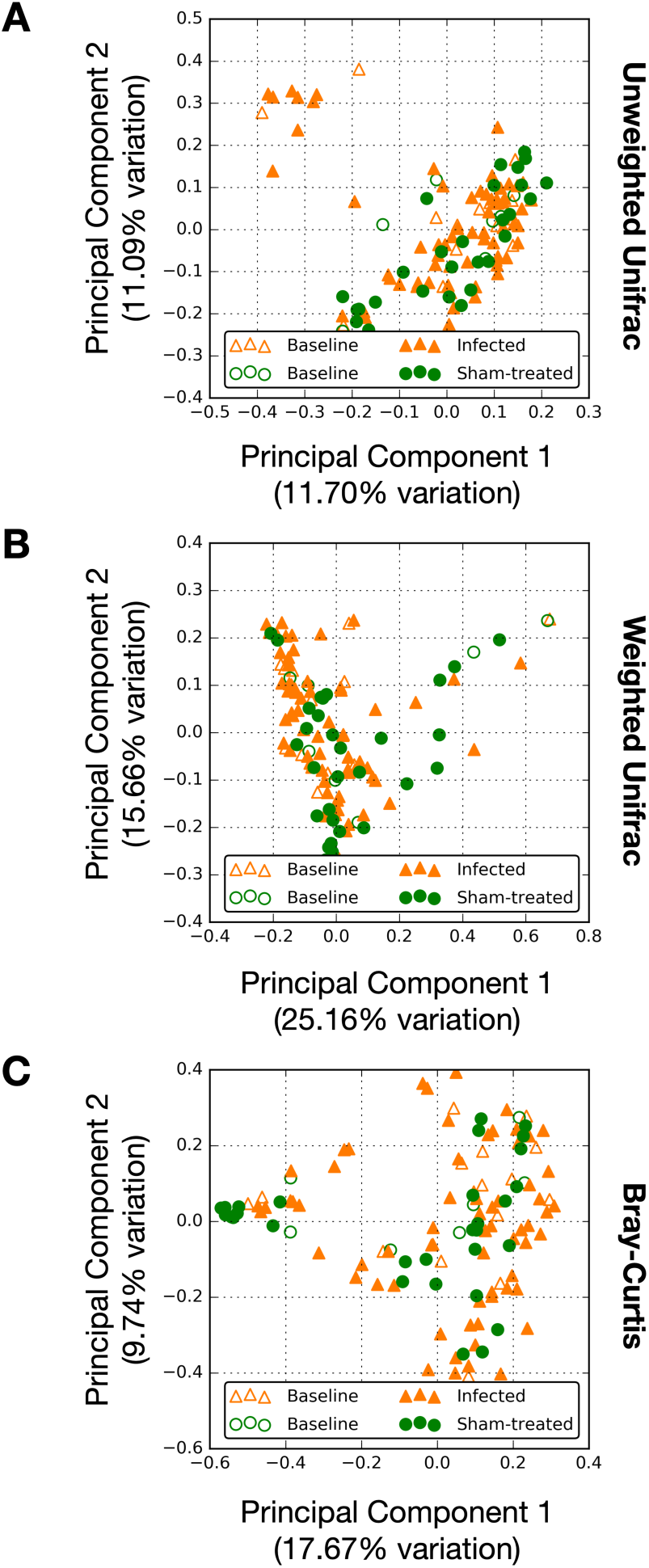
Principal coordinate analysis of nasal microbiota. The similarities of temporal nasal microbiota were projected onto a two-dimensional space. Shown are projections made using the unweighted Unifrac (**A**; p=0.305) and weighted Unifrac (**B**; p=0.301) distances, as well as Bray-Curtis dissimilarity (**C**; p=0.337) colored by infection status. Reported p-values are calculated using non-parametric multivariate analysis of variance with Infection as a main effect. Sample coloring by subject and time-point are provided in Figure S2.

Multiple statistical analyses quantitatively supported the conclusion that host phenotypes, aside from subject identity, are weak drivers of microbiota variation in our dataset. Non-parametric comparisons based on Monte Carlo permutations of unweighted and weighted Unifrac distances, as well as Bray-Curtis dissimilarity, revealed significantly higher inter-individual subject distances than intra-individual subject distances across the entire subject pool and when testing the sham-treated and infected cohorts separately (P<0.001; student’s two-sample t-test). Furthermore, semi-parametric Analysis of Variance (PERMANOVA) showed that subject identity had a significant association with microbiota composition (R^2^=0.457; p<0.001; Table 2). Infection status was not significantly associated with community structure (p=0.338, Table 2) nor when interacting with time (R^2^=0.033; p=0.601; Table 2). Time as a singular effect was not found to be deterministic of microbial structure (R^2^=0.046; p=0.113; Table 2).

**Table 2:**
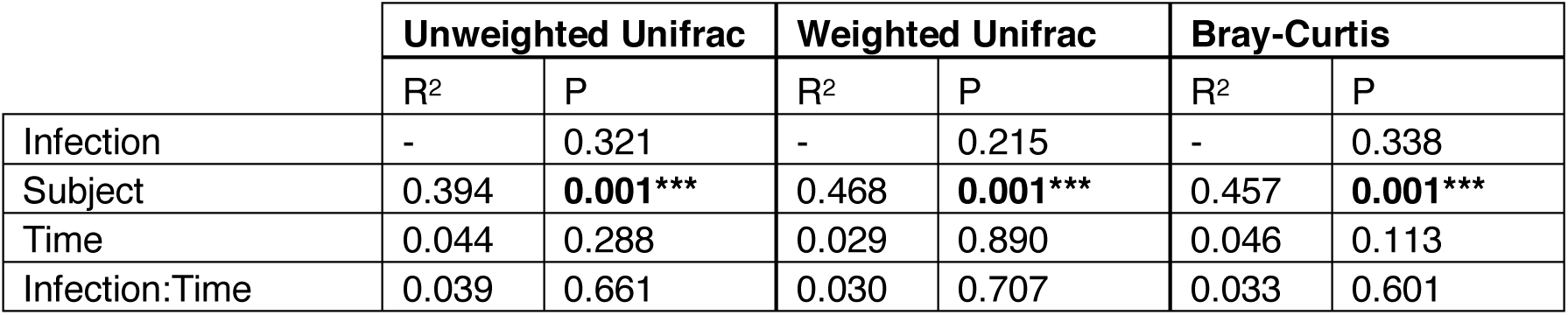
Results of variation partition testing. Analysis of variance was carried out using non-parametric multivariate ANOVA (*Methods*) applied to three different measures of β-diversity. R-squared values are provided when computed by the statistical package used to test a given effect.

We investigated whether specific microbial taxa could be associated with HRV infection. The most abundant phyla were Firmicutes (median=40.37%, MAD=10.30%), Actinobacteria (median=21.21%, MAD=10.27%), and Proteobacteria (median=21.33%, MAD=11.08%) across all subjects. We observed no significant differences between levels of phyla between sham-treated subjects and the infected cohort after FDR correction (p>0.05; Wilcoxon rank-sum tests; Figure 4, Table S4). We also did not observe changes in the relative levels of dominant taxa when samples from sham-treated and infected subjects were broken down by time point: before (t=-24 hours), during (t=8h to 96h), and after (t=120h) HRV infection (p>0.05; Kruskal-Wallis H tests; Table S5A). We then repeated the prior statistical tests at the genus level (the finest taxonomic level at which 16S rRNA are generally assigned with confidence with only a minimal loss of taxonomic resolution [37]). Again, no changes in the relative levels of dominant genera when grouping the infected and sham-treated cohorts by time point were observed (p>0.05; Kruskal-Wallis H tests; Table S5B). Comparison of genera in the sham-treated and infected cohorts with abundance greater than 1% during infection revealed no genera to be significantly associated with HRV infection after FDR correction (p>0.05; Wilcoxon rank-sum tests; Table S6).

**Figure 4.**
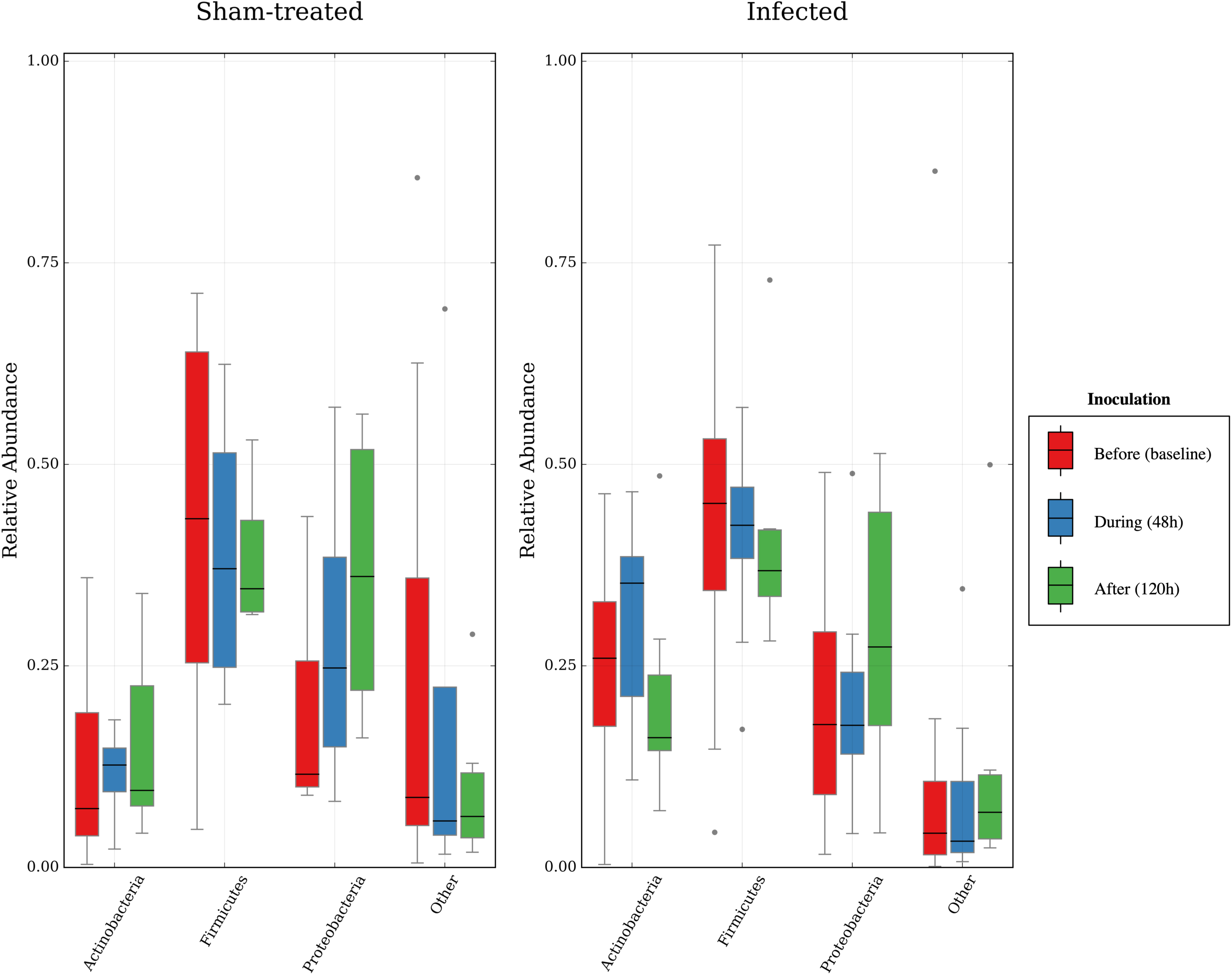
Box and whisker plot of relative abundance of dominant phyla by time point. Counts for sham-treated samples (before: n=7, during: n=4, after: n=6). Counts for infected samples (before: n=15, during: n=12, after: n=10). No significant differences during infection among all phyla were observed (p>0.05; Wilcoxon rank-sum).

Since significant bacterial dynamics could be obscured by the high number of taxonomic groups we tested for association with infection, we also performed a genus-level clustering analysis (Figure 5A). This analysis was designed to group genera with shared dynamics, easing microbiota interpretation and reducing the number hypotheses tested in statistical analyses [19, 34]. Clustering showed six major groups of bacterial taxa which accounted for at least 1% subjects’ total median abundance at one or more time points (Figure 5B/C). We did not observe significant differences in the abundance of these major clusters at any time point among HRV patients relative to subjects in the sham-treated cohort (p>0.05; Wilcoxon rank-sum tests; Table S7).

**Figure 5.**
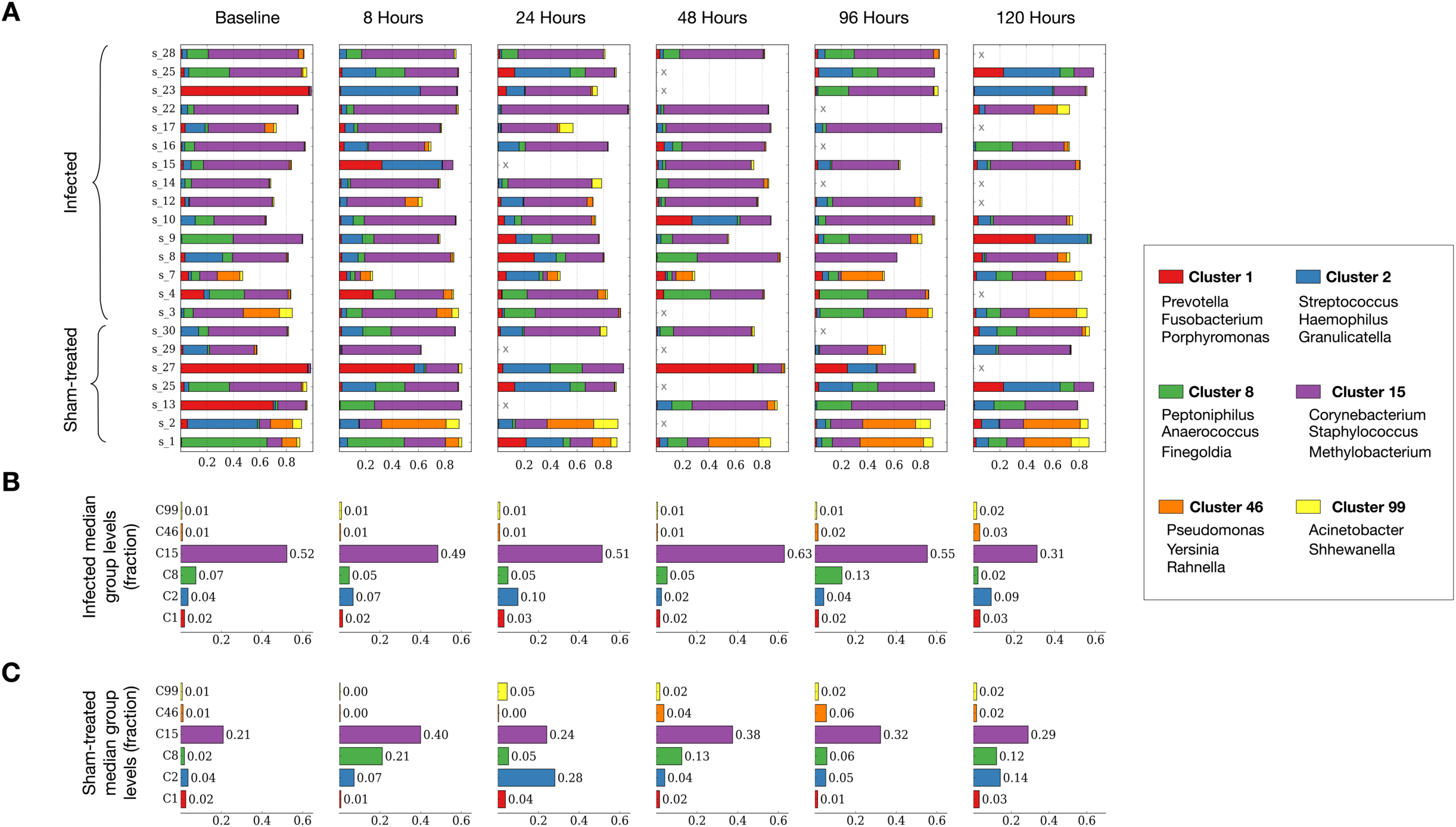
Abundance of nasal microbiota from the sham-treated and infected cohorts over time. (**A**) To simplify analysis, highly correlated genera were grouped according to their dynamics. Microbiota from sham-treated subjects are shown in green. Absent samples are shown with an X. (**B**) Median group abundances across infected subjects. (**C**) Median group abundances across sham-treated subjects. Abundance values are shown next to the bars (p > 0.05; Wilcoxon rank-sum). The three most abundant genera in each cluster are shown in the legend above. Genera assignments to clusters not shown can be found in Table S9.

Finally, we hypothesized that taxonomic dynamics within subjects’ microbiota might be more strongly associated with immune response than infection status. It has been previously demonstrated during viral infection that commensal bacteria augment immunity and antiviral gene expression [38]. As a result, we looked to identify statistically significant microbial-leukocyte relationships that may exist in the URT. The differential relative abundance of the six major clusters of genera at every timepoint was correlated with differences in counts of white blood cells, neutrophils, lymphocytes, monocytes, eosinophils, and basophils at the corresponding timepoints. No significant relationships were found between these bacterial and immune variables (Spearman’s rank correlation coefficient tests; p>0.05; Table S8).

## DISCUSSION

Here, we used HRV challenge to examine if and how nasal microbiota respond to viral infection. We found a lack of evidence for orderly dynamics and measurable differences in the microbiota of the nasopharynx following perturbation. HRV infection was not associated with changes in community diversity nor associated with changes in community structure or the levels of specific bacterial taxa. Within the infected cohort we found no differences in diversity associated with symptom severity. Furthermore, we were unable to relate any specific taxa with a subject’s susceptibility to HRV infection. No associations emerged between bacterial groups and host phenotypes, like infection status, or between microbiota variation and time past inoculation. Additionally, no correlations were found between leukocyte levels and bacterial genera. Subject identity was the strongest driver of overall community structure identified in our dataset.

Interpreting our lack of evidence for a URT microbiota response to HRV infection invites consideration of our samples size and power to detect meaningful effects. Determining an appropriate sample size for a microbiota study ultimately depends on the strength of an expected effect [39]. Here, we were motivated to examine the effects of HRV infection on URT microbiota in part because prior investigations of other pathogens in the nose and throat had revealed striking dynamics among commensal microbiota. We note that orderly changes in the abundance of bacterial taxa following infection by pathogens like *Vibrio cholera* [40], *Clostridium difficile* [41] and *Salmonella* [42] have also been observed in the guts of individuals or animals. We can estimate our likelihood of detecting such changes by considering one of our prior studies, in which we observed a predictable succession of microbiota following pathogen colonization in the human gut [19]. We found that levels of a common microbial genus in healthy fecal samples (*Prevotella, μ*=36%, *σ*=19%) were reduced among 10 infected individuals (*μ*=6%, *σ* =14%), and that overall microbiota Shannon diversity was also lower among individuals with cholera (*μ*=2.88, *σ*=0.96) than in healthy subjects (*μ*=4.82, *σ*=0.82). Power analyses indicate that should post-infection taxonomic or diversity responses among URT microbiota have similar effect sizes, we would have had a 92-98% chance of detecting such effects. Therefore, given our lack of significant statistical findings here, we conclude that our HRV challenge cohort likely did not experience an URT microbiota response of similar intensity and inter-individual consistency as in prior microbiota studies of infection. Ultimately, should reproducible statistical associations exist between our HRV infection model and URT microbiota, their effect sizes may be modest and require hundreds of human samples to detect [43].

The lack of a clear URT microbiota response to HRV infection presented here contrasts with prior reports that HRV infection was associated with significant changes in levels of select genera. Differences between this study and prior ones include differences in cohort size and broader age demographic of this study population as compared to earlier reports [23, 44]. Also, discrepancies in sampling methods and location across studies may influence our ability to observe patterns. For example, the microbiota measured in our nasal lavages represents bacteria sampled across multiple, and likely heterogenous, locations in the URT [45]. Still, none of the bacterial taxa previously associated with HRV were conserved across the prior studies and only two studies, Allen et al. and Hofstra et al., were explicit in their control for multiple hypothesis testing across bacterial taxa. Moreover, our study is consistent with prior HRV challenge reports in the lack of a reproducible response to HRV. Only one of the prior studies reported decreases in URT alpha-diversity during HRV infection [23], and none detected statistically significant shifts in overall community structure (*e*.*g*. via PERMANOVA) following HRV infection. Ultimately, understanding why viral infection does not always affect resident microbiota is likely to be an informative area of future research, as modulation of the host immune response by viruses is a complex and burgeoning field [46]. Knowing if and why viral infection does not alter microbiota will help define the limits of cross-talk between viral and bacterial immune responses.

The resistance of human URT microbiota to HRV infection also has implications for the pathogenesis of respiratory co-infection. Primary viral infections in the respiratory tract can be exploited by opportunistic bacterial pathogens [47], and a retrospective analysis of the 1918-1919 influenza pandemic suggests multiple waves of succession following infection resulted in secondary bacterial pneumonia [48]. Our finding that URT microbiota lack a consistent response to HRV infection suggests that if bacterial pathogens do exploit HRV infection, they do so opportunistically. No one bacterial species or group appears favored to grow in the wake of HRV infection.

## Conclusions

Among our study participants, we did not identify significant associations between HRV and human URT microbiota. The severity of clinical symptoms was independent of community diversity. We also did not identify host immune markers that were correlated with URT microbiota composition among the sham-treated and infected cohorts. Subject identity was found to be the strongest determinant of URT microbial community structure in our dataset. Our findings support the hypothesis that the nasopharyngeal microbial community is individualized and do not reveal stereotypical microbiota responses to rhinovirus challenge.

## Data Availability

A portion of data is available in the linked supplementary information. All other data that supports the findings of this study are available from the corresponding author(s) upon reasonable request.

## Acknowledgements

We would like to thank Stephen Brewer and Sunil Suchindran for statistical expertise and study volunteers for their participation. This work used a high-performance computing facility partially supported by grant 2016-IDG-1013 (“HARDAC+: Reproducible HPC for Next-generation Genomics”) from the North Carolina Biotechnology Center. L.A.D. acknowledges support from the Alfred P. Sloan Research Fellowship. S.N.N acknowledges support from the Duke Institute for Genomic Sciences and Policy summer fellowship.

## SUPPLEMENTARY INFORMATION

**Figure S1:**
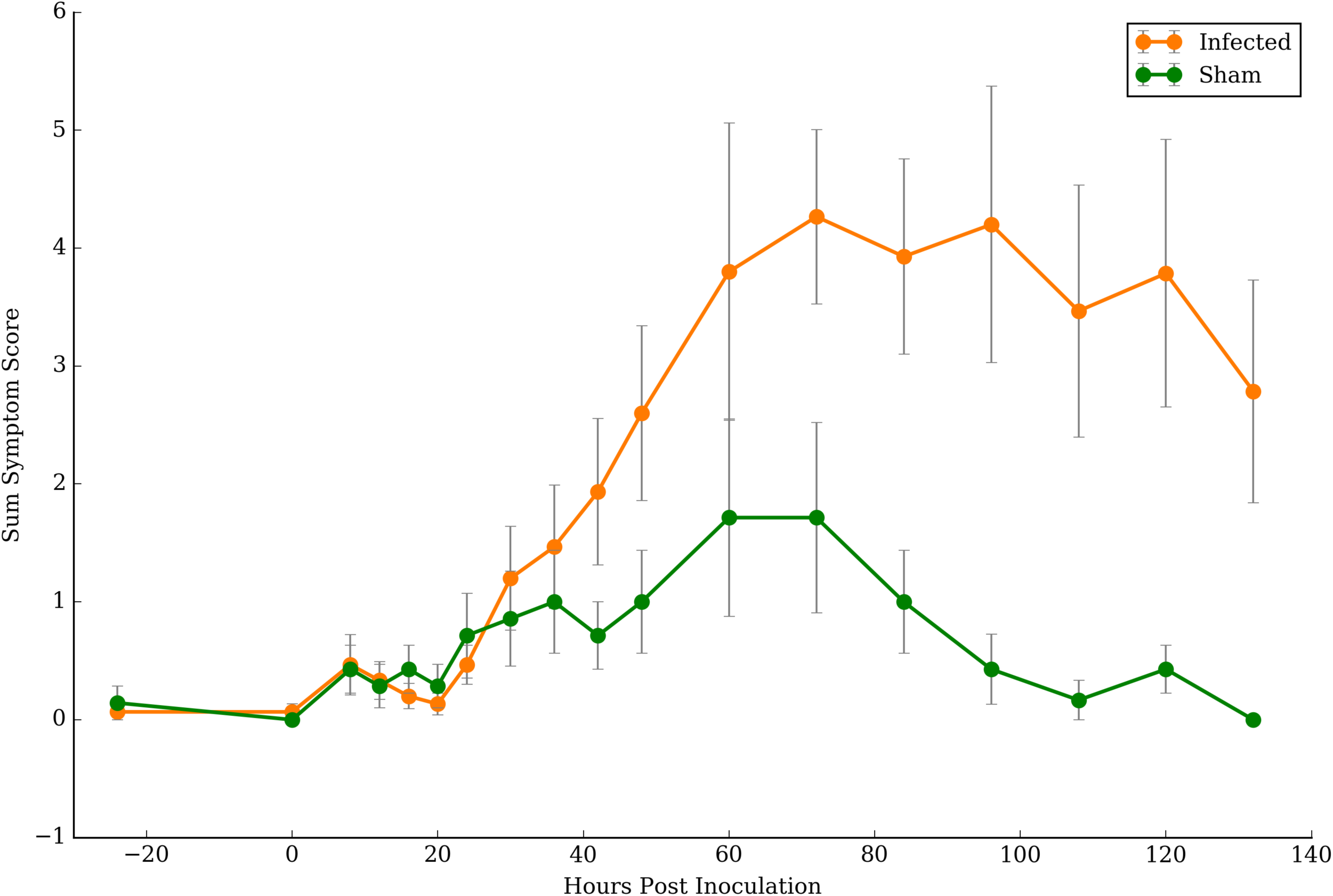
Mean symptom scores of sham and infected cohorts. Symptom scores were collected periodically throughout a week period. Error bars represent the standard error of the mean. The symptom scale for the categories of runny nose, stuffy nose, sneezing, coughing, malaise, sore throat, fever, headache, shortness of breath, and earaches was defined as 0-none/unknown, 1-mild, 2-moderate, 3-severe, 4-very severe.

**Figure S2:**
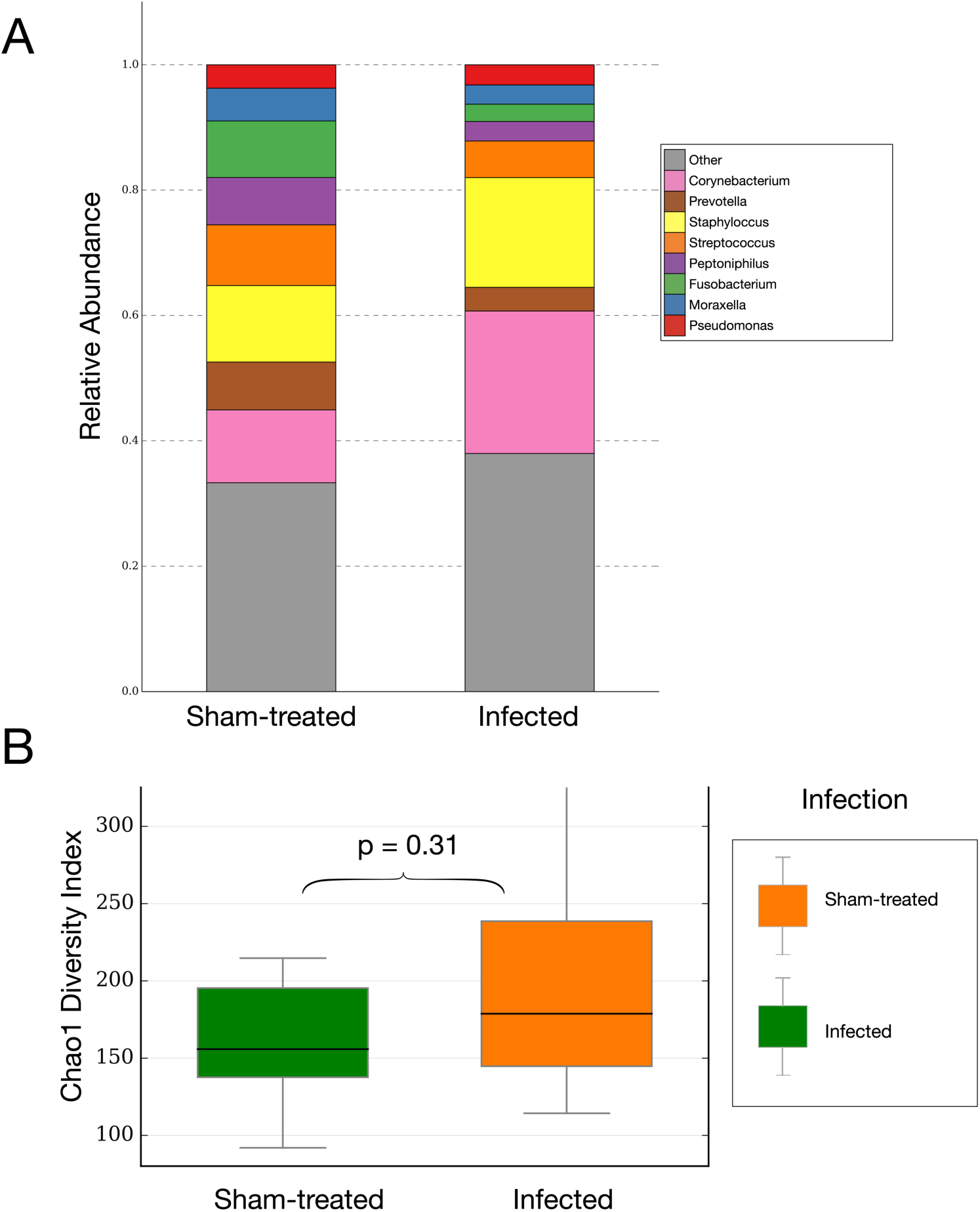
Baseline taxonomic composition of sham-treated and infected cohorts. (**A**) Stacked bar chart of the median abundance of most abundant genera within the two cohorts during infection at baseline (t=-24h). (**B**) Box and whisker plot of Chao1 diversity index. Sham-treated (n=7) versus infected (n=15). No significant differences in alpha diversity between the two cohorts were found (p > 0.05; Wilcoxon rank-sum) (see figure S3 for rarefaction curve).

**Figure S3:**
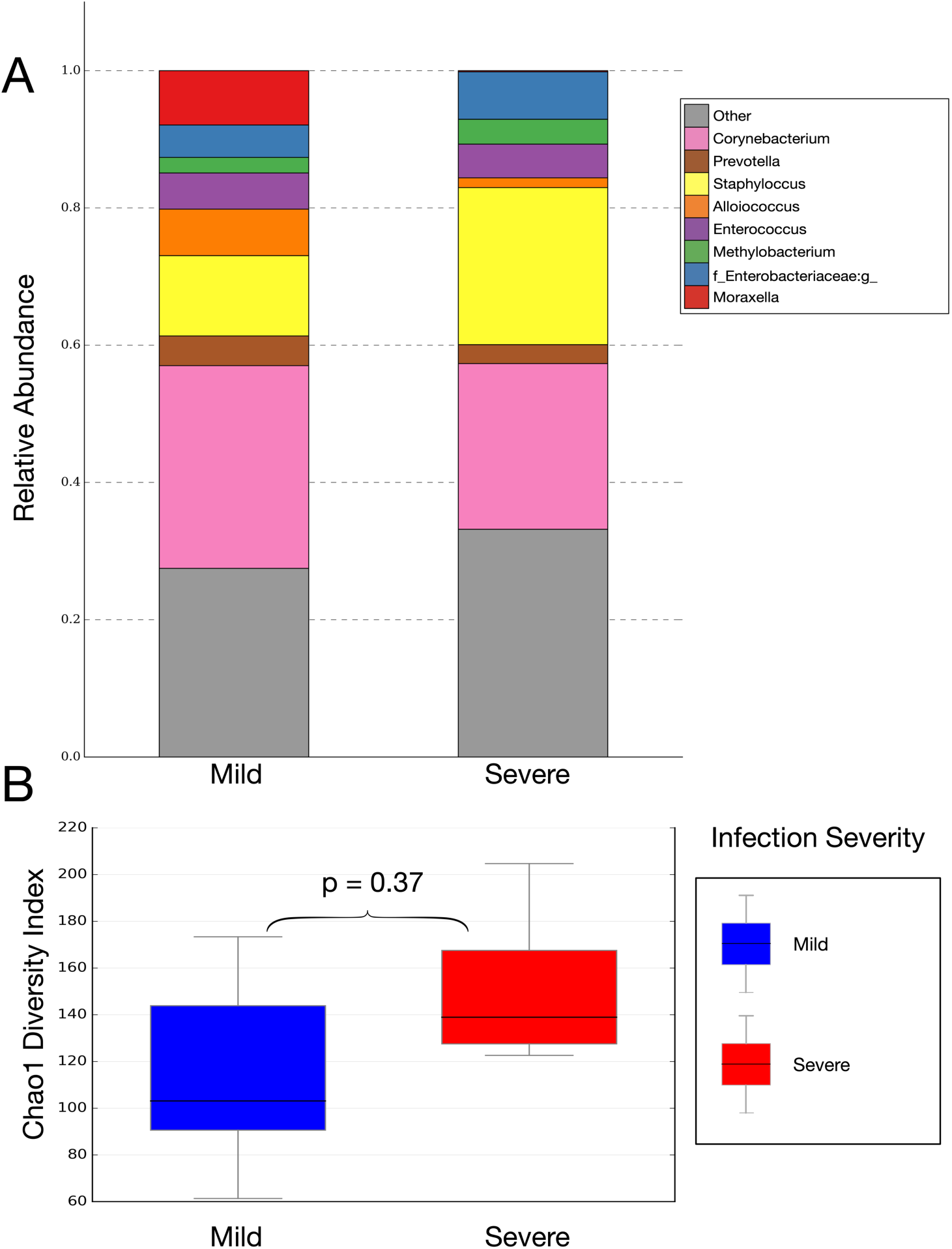
Composition of Mild and Severe HRV response groups. **A**) Stacked bar chart of the median abundance of most abundant genera within the two cohorts during infection at t=48h (p > 0.05; Wilcoxon rank-sum). (**B**) Box and whisker plot of Chao1 diversity index. Response severity as determined by symptom score: Mild (n=7) vs Severe (n=8). No significant differences in alpha diversity between the two groups were found (p > 0.05; Wilcoxon rank-sum).

**Figure S4.**
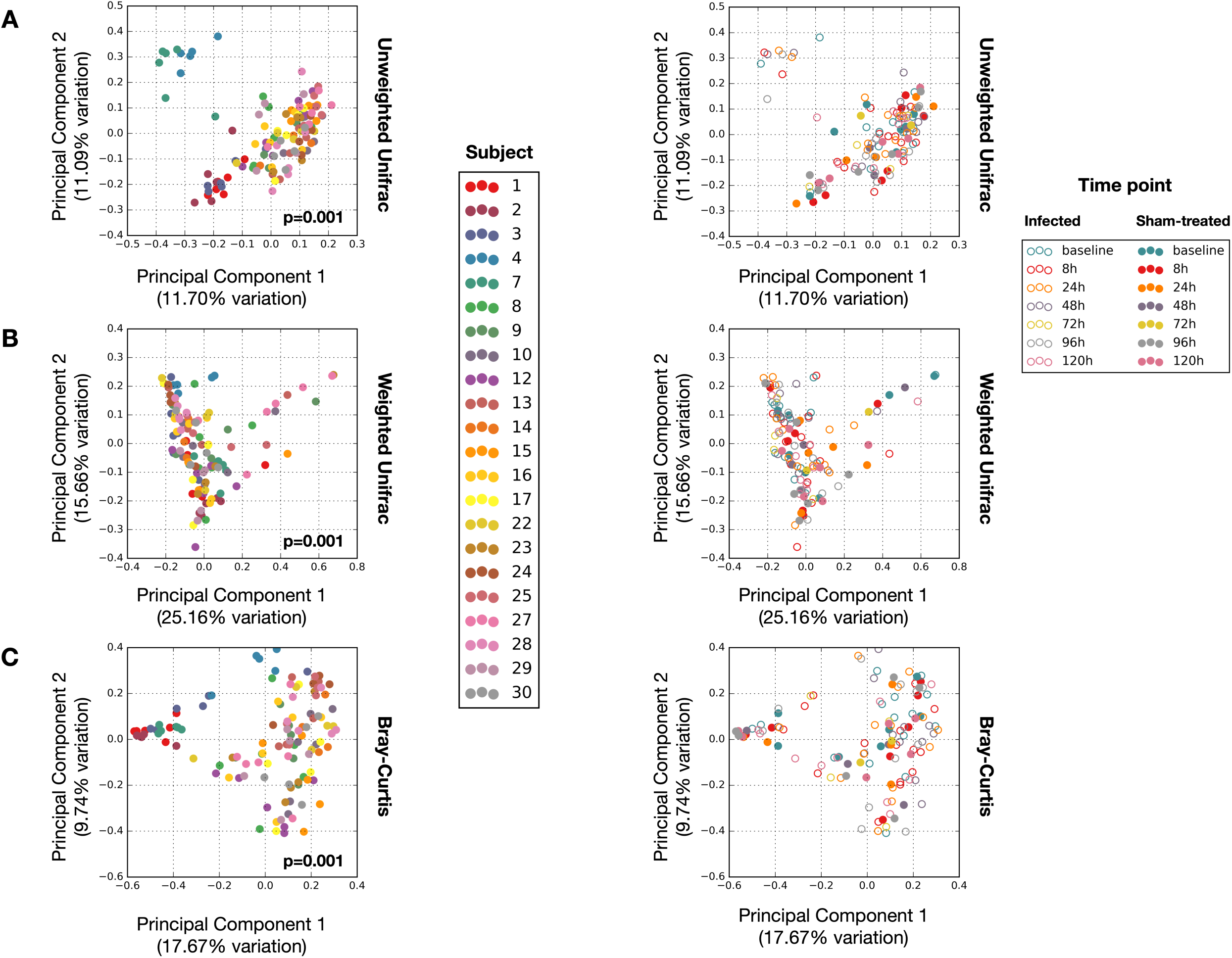
Principal coordinate analysis of nasal microbiota. Projections were made using the unweighted Unifrac (**A**) and weighted Unifrac (**B**) distances, as well as the Bray-Curtis dissimilarity (**C**). Samples are colored by individual (left column) and time (right column).

**Figure S5:**
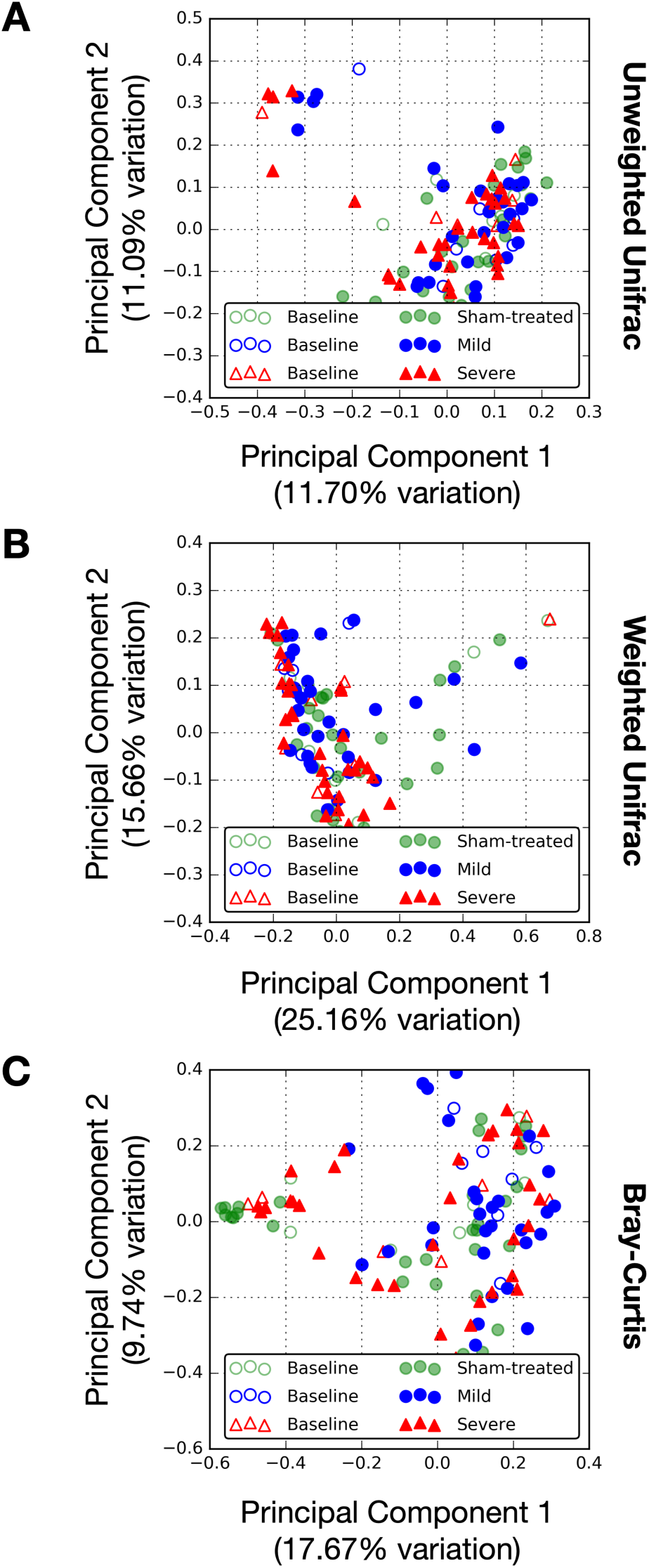
Principal coordinate analysis of nasal microbiota within the infected cohort. Shown are projections made using the unweighted Unifrac, weighted Unifrac distances, and Bray-Curtis dissimilarity colored by infection response severity and health status.

**Figure S6.**
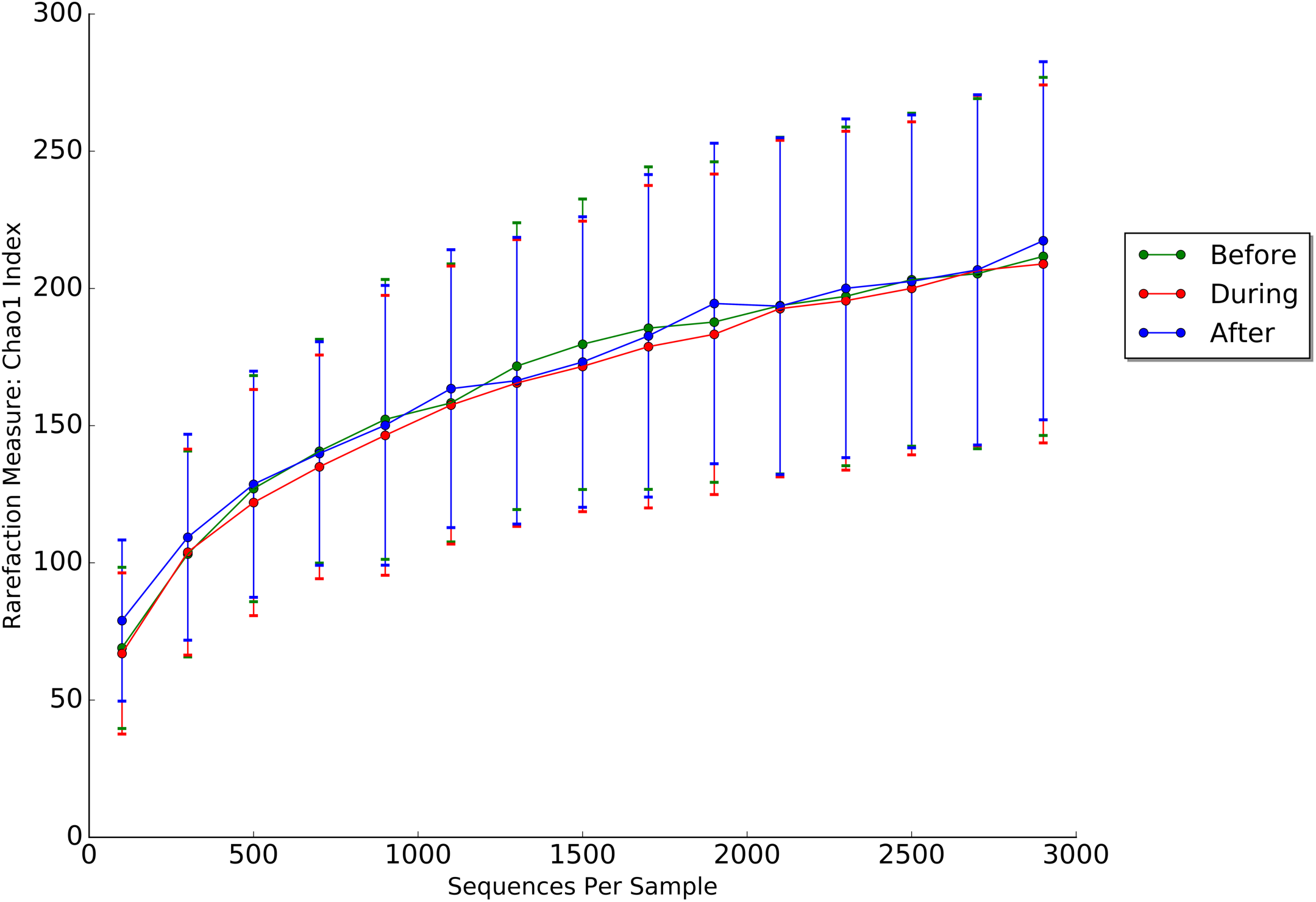
Alpha diversity rarefaction curve demonstrating the relationship between sample size and the number of observed species in infected samples. Lines are labeled by time following inoculation (before: t=-24 hours, during: t=8h to 96h, after: t=120h). When comparing the sham-treated and infected cohorts, no significant differences in sequences per sample were found (p>0.05, Wilcoxon rank-sum; Table S10).

**Table S1: Raw reported symptom scores**.

**Table S2: Complete blood count sample collections**. Subjects had samples taken 24 hours to inoculation with virus (baseline) and at set intervals following HRV challenge.

**Table S3: Subject assignment to mild and severe infection groups**. The highest symptom score reported in the categories of runny nose, stuffy nose, sneezing, cough, malaise, sore throat, headache, shortness of breath, and earache on each day was summed over the course of the week and is reported below. The median of this metric serves as the cutoff for mild (n=7) and severe (n=8) response HRV groups.

**Table S4: Wilcoxon rank-sum test** to test the null hypothesis that the relative abundances of phyla were the same between infected and healthy individuals during infection (t=48h).

**Table S5A: Kruskal-Wallis H test of dominant phyla** to test the null hypothesis that the relative abundances of phyla were the same across time points in healthy and infected subjects.

**Table S5B: Kruskal-Wallis H test of dominant genera** to test the null hypothesis that the relative abundances of the dominant genera were the same across time points in healthy and infected subjects.

**Table S6: Wilcoxon rank-sum test** to test the null hypothesis that the relative abundances of genera with median abundance greater than 1% during infection (t=48h) were the same between infected and healthy individuals.

**Table S7: Comparison of clusters classified as dominant during health and infection (t=48h)**.

**Table S8: Results of Spearman Rank Order Correlation**. R: Spearman correlation coefficient. P: FDR-corrected p-value (n=75).

**Table S9: Genera assignment to clusters**.

**Table S10: Wilcoxon rank-sum test to test the null hypothesis that the sequences per sample were the same between infected and sham samples**.

## BIBLIOGRAPHY

1. Faith JJ, Guruge JL, Charbonneau M, et al. The long-term stability of the human gut microbiota. Science 2013; 341:1237439.

2. David LA, Materna AC, Friedman J, et al. Host lifestyle affects human microbiota on daily timescales. Genome biology 2014; 15:R89.

3. Oh J, Byrd AL, Park M, Program NCS, Kong HH, Segre JA. Temporal Stability of the Human Skin Microbiome. Cell 2016; 165:854–66.

4. Stecher B, Robbiani R, Walker AW, et al. Salmonella enterica serovar typhimurium exploits inflammation to compete with the intestinal microbiota. PLoS biology 2007; 5:2177–89.

5. Iwase T, Uehara Y, Shinji H, et al. Staphylococcus epidermidis Esp inhibits Staphylococcus aureus biofilm formation and nasal colonization. Nature 2010; 465:346.

6. Man WH, de Steenhuijsen Piters WAA, Bogaert D. The microbiota of the respiratory tract: gatekeeper to respiratory health. Nature Reviews Microbiology 2017; 15:259.

7. Lemon KP, Klepac-Ceraj V, Schiffer HK, Brodie EL, Lynch SV, Kolter R. Comparative analyses of the bacterial microbiota of the human nostril and oropharynx. mBio 2010; 1:e00129–10.

8. Whelan FJ, Verschoor CP, Stearns JC, et al. The loss of topography in the microbial communities of the upper respiratory tract in the elderly. Annals of the American Thoracic Society 2014; 11:513–21.

9. Biesbroek G, Tsivtsivadze E, Sanders EA, et al. Early respiratory microbiota composition determines bacterial succession patterns and respiratory health in children. American journal of respiratory and critical care medicine 2014; 190:1283–92.

10. Teo SM, Tang HH, Mok D, et al. Airway microbiota dynamics uncover a critical window for interplay of pathogenic bacteria and allergy in childhood respiratory disease. Cell host & microbe 2018; 24:341-52. e5.

11. Ding T, Song T, Zhou B, et al. Microbial composition of the human nasopharynx varies according to influenza virus type and vaccination status. mBio 2019; 10:e01296–19.

12. Tarabichi Y, Li K, Hu S, et al. The administration of intranasal live attenuated influenza vaccine induces changes in the nasal microbiota and nasal epithelium gene expression profiles. Microbiome 2015; 3:74.

13. Salk HM, Simon WL, Lambert ND, et al. Taxa of the Nasal Microbiome Are Associated with Influenza-Specific IgA Response to Live Attenuated Influenza Vaccine. PLOS ONE 2016; 11:e0162803.

14. Edouard S, Million M, Bachar D, et al. The nasopharyngeal microbiota in patients with viral respiratory tract infections is enriched in bacterial pathogens. European Journal of Clinical Microbiology & Infectious Diseases 2018; 37:1725–33.

15. Langevin S, Pichon M, Smith E, et al. Early nasopharyngeal microbial signature associated with severe influenza in children: a retrospective pilot study. Journal of General Virology 2017; 98:2425–37.

16. de Steenhuijsen Piters WA, Heinonen S, Hasrat R, et al. Nasopharyngeal microbiota, host transcriptome, and disease severity in children with respiratory syncytial virus infection. American journal of respiratory and critical care medicine 2016; 194:1104–15.

17. Ederveen TH, Ferwerda G, Ahout IM, et al. Haemophilus is overrepresented in the nasopharynx of infants hospitalized with RSV infection and associated with increased viral load and enhanced mucosal CXCL8 responses. Microbiome 2018; 6:10.

18. Sonawane AR, Tian L, Chu C-Y, et al. Microbiome-transcriptome interactions related to severity of respiratory syncytial virus infection. Scientific reports 2019; 9:1–14.

19. David LA, Weil A, Ryan ET, et al. Gut microbial succession follows acute secretory diarrhea in humans. MBio 2015; 6:e00381–15.

20. Dubourg G, Edouard S, Raoult D. Relationship between nasopharyngeal microbiota and patient’s susceptibility to viral infection. Expert review of anti-infective therapy 2019; 17:437–47.

21. Rosas-Salazar C, Shilts MH, Tovchigrechko A, et al. Nasopharyngeal Microbiome in Respiratory Syncytial Virus Resembles Profile Associated with Increased Childhood Asthma Risk. American Journal of Respiratory and Critical Care Medicine 2016; 193:1180–3.

22. Hofstra JJ, Matamoros S, van de Pol MA, et al. Changes in microbiota during experimental human Rhinovirus infection. BMC Infect Dis 2015; 15:336.

23. Allen EK, Koeppel AF, Hendley JO, Turner SD, Winther B, Sale MM. Characterization of the nasopharyngeal microbiota in health and during rhinovirus challenge. Microbiome 2014; 2:22.

24. Zaas AK, Chen M, Varkey J, et al. Gene expression signatures diagnose influenza and other symptomatic respiratory viral infections in humans. Cell Host Microbe 2009; 6:207–17.

25. Turner RB, Weingand KW, Yeh CH, Leedy DW. Association between interleukin-8 concentration in nasal secretions and severity of symptoms of experimental rhinovirus colds. Clin Infect Dis 1998; 26:840–6.

26. Jackson GG, Dowling HF, Spiesman IG, Boand AV. Transmission of the common cold to volunteers under controlled conditions. I. The common cold as a clinical entity. AMA Arch Intern Med 1958; 101:267–78.

27. Turner RB. Ineffectiveness of intranasal zinc gluconate for prevention of experimental rhinovirus colds. Clin Infect Dis 2001; 33:1865–70.

28. Caporaso JG, Lauber CL, Walters WA, et al. Global patterns of 16S rRNA diversity at a depth of millions of sequences per sample. Proceedings of the National Academy of Sciences 2011; 108:4516–22.

29. Caporaso JG, Lauber CL, Walters WA, et al. Ultra-high-throughput microbial community analysis on the Illumina HiSeq and MiSeq platforms. The ISME Journal 2012; 6:1621–4.

30. DeSantis TZ, Hugenholtz P, Larsen N, et al. Greengenes, a chimera-checked 16S rRNA gene database and workbench compatible with ARB. Appl Environ Microbiol 2006; 72:5069–72.

31. Caporaso JG, Kuczynski J, Stombaugh J, et al. QIIME allows analysis of high-throughput community sequencing data. Nature methods 2010; 7:335–6.

32. Kuczynski J, Stombaugh J, Walters WA, González A, Caporaso JG, Knight R. Using QIIME to analyze 16S rRNA gene sequences from microbial communities. Curr Protoc Bioinformatics 2011; Chapter 10:Unit10.7-.7.

33. Oksanen J. Multivariate analysis of ecological communities in R: vegan tutorial. 2013.

34. David LA, Maurice CF, Carmody RN, et al. Diet rapidly and reproducibly alters the human gut microbiome. Nature 2014; 505:559–63.

35. Friedman J, Alm EJ. Inferring correlation networks from genomic survey data. PLoS computational biology 2012; 8:e1002687.

36. Jones E, Oliphant T, Peterson P. SciPy: Open source scientific tools for python. Available at: http://www.scipy.org.

37. Mizrahi-Man O, Davenport ER, Gilad Y. Taxonomic Classification of Bacterial 16S rRNA Genes Using Short Sequencing Reads: Evaluation of Effective Study Designs. PLOS ONE 2013; 8:e53608.

38. Ichinohe T, Pang IK, Kumamoto Y, et al. Microbiota regulates immune defense against respiratory tract influenza A virus infection. Proceedings of the National Academy of Sciences 2011; 108:5354–9.

39. Silverman JD, Shenhav L, Halperin E, Mukherjee S, David LA. Statistical Considerations in the Design and Analysis of Longitudinal Microbiome Studies. bioRxiv 2018:448332.

40. Hsiao A, Ahmed AS, Subramanian S, et al. Members of the human gut microbiota involved in recovery from Vibrio cholerae infection. Nature 2014; 515:423–6.

41. Buffie CG, Jarchum I, Equinda M, et al. Profound alterations of intestinal microbiota following a single dose of clindamycin results in sustained susceptibility to Clostridium difficile-induced colitis. Infection and immunity 2012; 80:62–73.

42. Bratburd JR, Keller C, Vivas E, et al. Gut Microbial and Metabolic Responses to Salmonella enterica Serovar Typhimurium and Candida albicans. mBio 2018; 9:e02032–18.

43. Sze MA, Schloss PD. Looking for a signal in the noise: revisiting obesity and the microbiome. MBio 2016; 7:e01018–16.

44. Kloepfer KM, Sarsani VK, Poroyko V, et al. Community-acquired rhinovirus infection is associated with changes in the airway microbiome. Journal of Allergy and Clinical Immunology 2017; 140:312-5. e8.

45. Proctor DM, Relman DA. The landscape ecology and microbiota of the human nose, mouth, and throat. Cell host & microbe 2017; 21:421–32.

46. Cadwell K. The virome in host health and disease. Immunity 2015; 42:805–13.

47. Ramphal R, Small PM, Shands JW, Fischlschweiger W, Small PA. Adherence of Pseudomonas aeruginosa to tracheal cells injured by influenza infection or by endotracheal intubation. Infection and immunity 1980; 27:614–9.

48. Morens DM, Taubenberger JK, Fauci AS. Predominant role of bacterial pneumonia as a cause of death in pandemic influenza: implications for pandemic influenza preparedness. J Infect Dis 2008; 198:962–70.

